# Post-acute sequelae of COVID-19 in a non-hospitalized cohort: results from the Arizona CoVHORT

**DOI:** 10.1101/2021.03.29.21254588

**Authors:** Melanie L Bell, Collin J Catalfamo, Leslie V. Farland, Kacey C Ernst, Elizabeth T Jacobs, Yann C Klimentidis, Megan Jehn, Kristen Pogreba-Brown

**Affiliations:** Department of Epidemiology and Biostatistics, Mel and Enid Zuckerman College of Public Health, The University of Arizona, Tucson, AZ, USA; School of Human Evolution and Social Change, Arizona State University, Tempe, AZ, USA

## Abstract

Clinical presentation, outcomes, and duration of COVID-19 has ranged dramatically. While some individuals recover quickly, others suffer from persistent symptoms, collectively known as post*-*acute sequelae of SAR-CoV-2 (PASC). Most PASC research has focused on hospitalized COVID-19 patients with moderate to severe disease. We used data from a diverse population-based cohort of Arizonans to estimate prevalence of various symptoms of PASC, defined as experiencing at least one symptom 30 days or longer. There were 303 non-hospitalized individuals with a positive lab-confirmed COVID-19 test who were followed for a median of 61 days (range 30-250). COVID-19 positive participants were mostly female (70%), non-Hispanic white (68%), and on average 44 years old. Prevalence of PASC at 30 days post-infection was 68.7% (95%CI 63.4, 73.9). The most common symptoms were fatigue (37.5%), shortness-of-breath (37.5%), brain fog (30.8%), and stress (30.8%). The median number of symptoms was 3 (range 1-20). Amongst 157 participants with longer follow-up (≥60 days), PASC prevalence was 77.1%.

## Introduction

Clinical presentation and outcomes of infection with SARS-CoV-2 range from asymptomatic infection to death. Duration of illness has also varied; some individuals recover quickly, but others experience persistent, often debilitating, symptoms. The experience of symptoms lasting 30 days and longer has been dubbed long COVID, or post*-*acute sequelae of COVID*-*19 (PASC). Most research to date has focused on PASC amongst hospitalized patients. However, most individuals with COVID-19 are not hospitalized, and PASC among these non-hospitalized individuals is poorly characterized. We aimed to describe the prevalence of post-COVID-19 symptoms amongst individuals who experienced mild-to-moderate COVID-19 using data from a diverse, population-based cohort.

## Methods

In May 2020, we began recruitment for a cohort study, CoVHORT, that aimed to investigate the impact of SARS-Cov-2 among Arizona residents (1). Briefly, targeted recruitment of confirmed COVID-19 participants occurred via case investigations for Arizona health departments conducted by The University of Arizona’s Student Aid for Field Epidemiology Response program (2). Recruitment from the greater community was conducted via invitations to participants of partnered COVID-19 studies, phased mailing campaigns, partnerships with community testing labs, and distribution of informational materials. Informed consent was obtained from all participants. Ethics approval was obtained from the University of Arizona Institutional Review Board (Protocol #2003521636A002).

The current study included participants who had a positive polymerase chain reaction (PCR) or antigen test for SARS-COV-2 and provided symptom data 30 days or longer following the positive test. We excluded 33 participants who reported that they were hospitalized. At 6-7 weeks post-report of incident COVID, participants were asked whether they were experiencing any of 25 new or recurring symptoms (an open field was included for symptoms not listed). We defined PASC as the presence of at least one symptom 30 days or longer following a positive test. Demographics, existing chronic conditions, disease severity, and symptom data were self-reported.

## Statistical Methods

Among COVID-19 positive participants, descriptive statistics were calculated by stratifying on PASC status, and compared using chi-square, t-tests, or non-parametric analogues. We calculated the prevalence and 95% confidence interval (CI) of PASC, number of symptoms, and self-reported COVID-19 related symptoms at ≥30, 30-59, and ≥ 60 days follow-up. We used Wald CIs for binary variables and distribution-free CIs for medians. We performed two sensitivity analyses. First, to account for potential dropout bias, where participants lost to follow-up may be less likely to experience PASC, we used multiple imputation (MI) with a delta adjustment, decreasing the likelihood of dropouts experiencing PASC by delta = 25%, 50%, and 75% (3). MI models included factors associated with dropout. Second, because stress is a symptom that many persons may be experiencing due to the pandemic, we quantified the number of individuals who experienced stress as their only symptom. All analyses were performed in SAS version 9.4 (Cary, NC).

## Results

Of the 3,468 participants in the CoVHORT as of 2/24/2021, 747 had a positive PCR or antigen test and were not hospitalized for their illness. Excluding participants who had incomplete COVID-19 testing information, 543 received a follow-up survey. Of these 543, 303 (55.8%) completed the follow-up surveys. Participants had a mean age of 44 years (range 12-82 years), were mostly female (70%), non-Hispanic white (68%), with college or greater education (38%), and with at least one pre-existing chronic condition (67%). The most commonly reported pre-existing conditions were seasonal allergies (42%), asthma (16%) and hypertension (15%). The overall mean self-reported severity rating of their COVID-19 illness was 4.6 out of 10. Individuals with PASC were more likely to have less education, at least one pre-existing chronic condition, seasonal allergies, and greater self-reported severity as compared to participants not experiencing PASC (Table 1). Females were more likely to experience PASC than males (73 versus 63%), however this did not reach the threshold of statistical significance (p=0.07).

**Table 1.** Characteristics of 303 Arizona CoVHORT participants with lab confirmed positive COVID-19 test, with 30 or more days of follow-up, from 28 May 2020 to 24 February 2021. Values are n (%) unless indicated otherwise.

| Characteristic | Total (n = 303) | PASC (n = 208) | No PASC (n = 95) | p-value<sup>a</sup> |
| --- | --- | --- | --- | --- |
| Follow-up (d), median (IQR) | 61 (50, 85) | 62 (53, 88) | 57 (48, 80) | 0.005 |
| Age (y), mean (SD) | 43.6 (16.6) | 43.8 (15.8) | 43.1 (18.2) | 0.73 |
| Female gender, n(%) | 212 (70.0) | 152 (73.1) | 60 (63.2) | 0.07 |
| Race/ethnicity |  |  |  | 0.70 |
| Hispanic | 69 (22.8) | 50 (24.0) | 19 (20.0) |  |
| Non-Hispanic White | 205 (67.7) | 139 (66.8) | 69 (72.6) |  |
| Other race | 19 (6.3) | 14 (6.7) | 5 (5.3) |  |
| Education |  |  |  | 0.0009 |
| Some college or less | 44 (14.5) | 39 (18.8) | 5 (5.3) |  |
| College or more | 115 (38.0) | 72 (34.6) | 43 (45.3) |  |
| Missing | 144 (47.5) | 97 (46.6) | 47 (49.5) |  |
| BMI (kg/m<sup>2</sup>), mean (SD) | 28.0 (7.0) | 28.5 (7.3) | 26.9 (6.2) | 0.08 |
| Currently smokes or vapes | 40 (12.7) | 27 (12.2) | 13 (13.7) | 0.72 |
| Any pre-existing chronic condition<sup>b</sup> | 203 (67.0) | 148 (71.2) | 55 (57.9) | 0.02 |
| Seasonal allergies | 127 (41.9) | 97 (46.6) | 30 (31.6) | 0.01 |
| Asthma | 48 (15.8) | 31 (14.9) | 17 (17.9) | 0.51 |
| Hypertension | 45 (14.9) | 34 (16.3) | 10 (10.5) | 0.28 |
| COVID-19 severity<sup>c</sup>, mean (SD) | 4.6 (2.4) | 5.1 (2.4) | 3.4 (1.9) | <0.0001 |

Percentages may not add to 100% due to rounding and missing data. BMI = body mass index. PASC = post-acute sequelae of COVID-19, defined as any symptom reported at ≥30 days follow-up.
<sup>a</sup> For the test of any reported symptom vs no reported symptom amongst non-missing values
<sup>b</sup> Self-reported pre-existing chronic conditions include: Thyroid disorder, asthma, chronic obstructive pulmonary disease, valley fever, emphysema, seasonal allergies, diabetes, pre-diabetes, cancer, heart disease, high blood pressure, liver disease, kidney disease, gastrointestinal conditions, depression/anxiety, autoimmune diseases.
<sup>c</sup> Self-reported COVID-19 Severity based on a scale from 1-10

At ≥30 days follow-up, 208 of 303 (68.7%) participants reported experiencing PASC. The median number of symptoms among people experiencing PASC was 3 (range 1-20), with a median follow-up of 63 days (range 30-250). The 10 most commonly reported symptoms among individuals with PASC at ≥30 days post-positive test fatigue (37.5%); shortness of breath (37.5%); brain fog (30.8%); stress (30.8%); altered smell or taste (26.4%); body aches or muscle pains (26.0%); insomnia (22.1%); headaches (20.7%); joint pain (20.2%); and congestion or runny nose (19.2%) (Table 2).

**Table 2.** Self-reported symptoms of COVID-19 positive participants experiencing PASC at 30 or more days of follow-up from 28 May 2020 - 24 February 2021 in Arizona.

|  | Follow-up ≥30 days |  | Follow-up 30-59 days |  | Follow-up ≥60 days |  |
| --- | --- | --- | --- | --- | --- | --- |
|  | N = 208 |  | N = 87 |  | N = 121 |  |
|  | Estimate | 95% CI | Estimate | 95% CI | Estimate | 95% CI |
| PASC, n(%) <sup>a</sup> | 208 (68.7) | 63.4, 73.9 | 87 (59.6) | 51.6, 67.5 | 121 (77.1) | 70.5, 83.6 |
| Number of symptoms, median (IQR) | 3 (1, 6) | 2, 4 | 3 (1, 5) | 2, 4 | 3 (2, 7) | 2, 4 |
| Follow-up (d), median (IQR) | 63 (53, 87.5) | 60.0, 68.6 | 51 (48, 55) | 49.9, 53.0 | 83 (68, 121) | 77.0, 91.0 |
| **Symptom, N (%)** |  |  |  |  |  |  |
| Fatigue | 78 (37.5) | 30.9, 44.1 | 21 (24.1) | 15.1, 33.1 | 57 (47.1) | 38.2, 56.0 |
| Shortness of breath | 78 (37.5) | 30.9, 44.1 | 23 (26.4) | 17.2, 35.7 | 55 (45.5) | 36.6, 54.3 |
| Confusion or brain fog | 64 (30.8) | 24.5, 37.0 | 24 (27.6) | 18.2, 37.0 | 40 (33.1) | 24.7, 41.4 |
| Stress | 64 (30.8) | 24.5, 37.0 | 23 (26.4) | 17.2, 35.7 | 41 (33.9) | 25.5, 42.3 |
| Changes in smell or taste | 55 (26.4) | 20.4, 32.4 | 25 (28.7) | 19.2, 38.2 | 30 (24.8) | 17.1, 32.5 |
| Body aches or muscle pains | 54 (26.0) | 20.0, 31.9 | 17 (19.5) | 11.2, 27.9 | 37 (30.6) | 22.4, 38.8 |
| Insomnia | 46 (22.1) | 16.5, 27.8 | 19 (21.8) | 13.2, 30.5 | 27 (22.3) | 14.9, 29.7 |
| Headaches | 43 (20.7) | 15.2, 26.2 | 17 (19.5) | 11.2, 27.9 | 26 (21.5) | 14.2, 28.8 |
| Joint pain | 42 (20.2) | 14.7, 25.6 | 15 (17.2) | 9.3, 25.2 | 27 (22.3) | 14.9, 29.7 |
| Congestion or runny nose | 40 (19.2) | 13.9, 24.6 | 21 (24.1) | 15.1, 33.1 | 19 (15.7) | 9.2, 22.2 |
| Gastro-intestinal <sup>b</sup> | 38 (18.3) | 13.0, 23.5 | 15 (17.2) | 9.3, 25.2 | 23 (19.0) | 12.0, 26.0 |
| Tachycardia | 36 (17.3) | 12.2, 22.4 | 13 (14.9) | 7.5, 22.4 | 23 (19.0) | 12.0, 26.0 |
| Cough | 32 (15.4) | 10.5, 20.3 | 11 (12.6) | 5.7, 19.6 | 21 (17.4) | 10.6, 24.1 |
| Rash/skin conditions<sup>d</sup> | 32 (15.4) | 10.5, 20.3 | 12 (13.8) | 6.5, 21.0 | 20 (16.5) | 9.9, 23.1 |
| Dizziness/lightheadedness | 26 (12.5) | 8.0, 17.0 | 10 (11.5) | 4.8, 18.2 | 16 (13.2) | 7.2, 19.3 |
| Chest pain or tightness | 31 (14.9) | 10.1, 19.7 | 13 (14.9) | 7.5, 22.4 | 18 (14.9) | 8.5, 21.2 |
| High blood pressure | 23 (11.1) | 6.8, 15.3 | 6 (6.9) | 1.6, 12.2 | 17 (14.1) | 7.9, 20.2 |
| Tinnitus | 19 (9.1) | 5.2, 13.0 | 6 (6.9) | 1.6, 12.2 | 13 (10.7) | 5.2, 16.3 |
| Menstrual issues<sup>c</sup> | 13 (8.6) | 4.6, 14.2 | 8 (12.5) | 5.6, 23.2 | 5 (5.7) | 1.9, 12.8 |
| Chills or sweats | 17 (8.2) | 4.5, 11.9 | 5 (5.8) | 0.9, 10.6 | 12 (9.9) | 4.6, 15.2 |
| Loss of appetite | 16 (7.7) | 4.1, 11.3 | 8 (9.2) | 3.1, 15.3 | 8 (6.6) | 2.2, 11.0 |
| Sore throat | 15 (7.2) | 3.7, 10.7 | 8 (9.2) | 3.1, 15.3 | 7 (5.8) | 1.6, 9.9 |
| Changes in vision | 13 (6.3) | 3.0, 9.5 | 4 (4.6) | 0.2, 9.0 | 9 (7.4) | 2.8, 12.1 |
| Pink eye | 9 (4.3) | 1.6, 7.1 | 1 (1.2) | 0.0, 3.4 | 8 (6.6) | 2.2, 11.0 |
| Fever | 8 (3.9) | 1.2, 6.5 | 1 (1.2) | 0.0, 3.4 | 7 (5.8) | 1.6, 9.9 |
| Hair loss | 5 (2.4) | 0.3, 4.5 | 0 (0.0) | 0.0, 0.0 | 5 (4.1) | 0.6, 7.7 |
| Other<sup>e</sup> | 2 (1.0) | 0.0, 2.3 | 0 (0.0) | 0.0, 0.0 | 2 (1.7) | 0.0, 3.9 |
| Erectile dysfunction | 1 (0.5) | 0.0, 1.4 | 0 (0.0) | 0.0, 0.0 | 1 (0.8) | 0.0, 2.4 |

Abbreviations: IQR = interquartile range, CI = confidence interval, PASC = post-acute sequelae of COVID-19, defined as any symptom reported at $\geq 30$ days follow-up.
<sup>a</sup> Denominator for PASC prevalence includes all COVID-19 positive participants with $\geq 30$ days of follow-up (n = 303), 30-59 days (n = 146), and $\geq 60$ days (n = 157)
<sup>b</sup> Diarrhea, constipation, abdominal pain, nausea, vomiting)
<sup>c</sup> Denominator includes women only (n = 152).
<sup>d</sup> COVID toes, hive-like rashes, skin discoloration, bruising
<sup>e</sup> Other includes write-in symptoms not classified

Among participants followed for 30-59 days post-diagnosis, 59.6% reported PASC and 77.1% at ≥60 days follow-up. Rates of symptoms were higher at longer follow-up; though, the most prevalent symptoms were similar: fatigue, shortness-of-breath, and brain fog. We found that only 6 participants reported stress as their sole symptom, demonstrating that our PASC prevalence is not driven by this non-specific symptom.

Compared to eligible participants with follow-up data, those without were younger (39 versus 44 years), more likely to be male (37 versus 30%), of Hispanic ethnicity (32 versus 23%), more likely to smoke or vape (25 versus 13%) and have lower education (60 versus 72% who had finished college). Disease severity rating was similar (4.9 versus 4.7 out of 10) as were rates of pre-existing conditions (64 versus 70%). In sensitivity analyses attempting to account for differences in follow-up using a not-at-random, MI model, we estimated a PASC prevalence of 69.5% (delta=0%), 62.2% (delta=25%), 54.8% (delta=50%) and 47.7% (delta=75%), where delta is the decrease in likelihood of PASC for imputed values.

## Discussion

We found among non-hospitalized lab confirmed COVID-19 positive participants, 68.7% experienced at least one symptom 30 days or longer past test-date. For individuals with ≥60 days follow-up, the prevalence of PASC was 73%. The most common symptoms were fatigue (37.5%), shortness of breath (37.5%), brain fog (30.8%), and stress (30.8%). We found that less education, having at least one pre-existing condition (in particular, seasonal allergies), and greater self-reported COVID-19 severity at enrollment were associated with higher prevalence of PASC. While higher prevalence of PASC was seen among women and smokers/vapers, these were not statistically significant. Notably, we did not observe different rates of PASC based on age or BMI.

Our estimated prevalence of PASC is only slightly less than those reported for hospitalized individuals: Huang et al. found a prevalence of 76% at 6 months(4); Carfi et al. reported PASC prevalence of 87% at 2 months(5). A systematic review of 7 studies of mixed follow-up and severity (hospitalized and non-hospitalized) by Lopez-Leon et al. found a PASC prevalence of 80%(6). Logue et al. and Sudre et al., however, reported a much lower prevalence of PASC of 33% among outpatients(7), and 13.3% in a mixed-severity group(8), respectively. Logue et al.’s prevalence of 33% could possibly be due to their longer follow-up of 169 days, as compared to our median follow-up time of 59 days(7). Their estimate of 33% is smaller than our most extreme sensitivity analysis (48%), where imputed values had a 75% reduced likelihood of experiencing PASC.

Our most common symptoms were similar to results from prior research; however, our prevalence of specific symptoms were lower than for hospitalized samples. Commonly reported symptoms across multiple studies have been fatigue, headache, attention disorder (presumably similar to our variable, brain fog), hair loss, shortness of breath, sleeping problems, joint pain, dyspnea, chest pain and loss of sense of smell or taste (3-6).

A strength of our study is that our sample is derived from non-hospitalized lab confirmed COVID-19 positive individuals recruited from public health surveillance efforts, a population in which PASC is not well-described. Our study also has limitations. Our response rate at follow-up was low (56%), and participants who completed follow-up questionnaires may differ from those who did not, possibly with a bias towards people who suffer from PASC. However, reported severity and having a pre-existing condition, both of which were associated with PASC, were similar between participants with and without follow-up. It is also possible that individuals suffering from severe PASC were too ill to complete follow-up surveys. We conducted a statistically principled sensitivity analysis for loss to follow-up, and still estimated a high prevalence of PASC (47.7% (delta=75%)). Our characterization of the PASC phenotype is limited to 25 symptoms; other researchers have included more symptoms in their assessment(9). Finally, we may have been underpowered to detect differences.

COVID-19 has infected more than 110,000,000 individuals worldwide as of 1 March 2021 (10). If 63% (the lower limit of our 95% CI) of survivors experience persistent symptoms, over 63,000,000 individuals could be affected by the long-term consequences of COVID-19. Our estimate at ≥60 days follow-up showed that 50% of our participants were suffering from of 3 or more symptoms, with 25% experiencing 7 or more symptoms. These figures portend a public health challenge of massive scale. Further research is needed to characterize the clinical spectrum of PASC more completely in a variety of populations, including investigating correlates of PASC, treatment options, and time to resolution.

## Data Availability

Due to privacy and ethical considerations, data are not publicly available.

## Author Contributions

MLB and CJC analyzed the data and wrote the first draft. KPG, LVF, KCE, ETJ, YCK and MJ made substantial contributions to designing the original study and acquiring the data. All authors contributed to revising the manuscript and take responsibility for its contents.

## Acknowledgements

We acknowledge the entire CoVHORT team and participants.

